# Some statistical theory for interpreting reference distributions

**DOI:** 10.1101/2024.07.23.24309680

**Authors:** Berk A. Alpay, John M. Higgins, Michael M. Desai

## Abstract

Reference distributions quantify the extremeness of clinical test results, typically relative to those of a healthy population. Intervals of these distributions are used in medical decision-making, but while there is much guidance for constructing them, the statistics of interpreting them for diagnosis have been less explored. Here we work directly in terms of the reference distribution, defining it as the likelihood in a posterior calculation of the probability of disease. We thereby identify assumptions of the conventional interpretation of reference distributions, criteria for combining tests, and considerations for personalizing interpretation of results from reference data. Theoretical reasoning supports that non-healthy variation be taken into account when possible, and that combining and personalizing tests call for careful statistical modeling.

---

Disease can perturb the abundances of analytes measurable by clinical laboratory technology. However, even when an analyte is physiologically expected to be associated with a disease, it is not immediately clear how to translate laboratory measurements to clinical decisions: how should numbers be mapped to actions [1, 2]?

Reference distributions, which represent the variation in test results among healthy people, provide one way to interpret clinical results on the assumption that it is noteworthy if a patient has an extreme enough value relative to the population [3]. Commonly, test results are displayed alongside reference intervals, thresholds of extremeness with respect to the reference distribution — a familiar sight to many who have had blood tests. One very large-scale United Kingdom study of more than half a million patients, for example, inferred a reference interval for albumin-adjusted serum calcium of 2.19 to 2.56 mmol/L [4]. Thus, if used in practice, test results outside this interval might be flagged. Indeed, the authors speculated that adopting this interval over the previous consensus interval of 2.20 to 2.60 mmol/L [5] would affect rates of calcium metabolism disorder diagnosis.

Studies of simulated and clinical data over several decades have informed guidelines [3, 6] about how to set these reference intervals. Topics of statistical recommendations include inferring intervals from data [7–9], subgrouping when it is inappropriate to compare a patient’s result against the entire population [10,11], and transferring intervals between laboratories [12].

As more data is gathered, it has been of increasing concern to create new references and tune existing ones. Reference intervals are reconsidered [13, 14], new ones are constructed [15], and sources of healthy variation are accounted for [16, 17] with an enduring goal being references personalized to the patient [18].

Although less abundant than studies on constructing intervals, important work has been done about how to interpret them for diagnosis [2, 19–21]. Because, in clinical practice, results are compared to reference intervals, previous theory has framed questions of interpretation by viewing reference intervals as thresholds of diagnosis. (Indeed, a reference distribution is generally seen as an intermediate object whose purpose is to yield a reference interval [6, 9, 22, 23].) It has been shown, critically, that the diagnostic predictiveness of intervals depends on the prevalence of the disease [19] and the distributions of results among both the healthy and non-healthy population [2, 20, 21]. However, it remains difficult to reason precisely about some diagnostic questions by approaching them directly in terms of true and false positives and negatives, even if these quantities are the valued endpoints.

Here, instead, we formalize the connection between reference values and the probability of disease in terms of distributions of results. This framework allows us to expressively analyze aspects of reference distributions and their resulting intervals. We identify assumptions of current clinical interpretation and reason about how to interpret reference distributions with respect to those of other analytes as well as features of each patient and the conditions of their test. Our analysis often complements and expands on prior work. We synthesize previous findings, recapitulate and counter various theoretical arguments, and highlight new considerations for constructing references. Our approach is strictly theoretical and intended to reason about interpreting reference distributions as generally as possible. Some of our example cases are extreme and may not have direct analogs in clinical data, although they are all intended to be useful for intuition of the underlying statistics.

## Reference distributions express likelihoods of results

Suppose a patient’s blood test reports a result for some analyte of, say, 0.25 mg/dL. How should this result be interpreted?

The test might have been taken to rule out or settle on some set of diagnoses, according to which additional tests will be performed or treatment will be adjusted. So, the test result *y* = 0.25mg/dL could be interpreted in terms of the probability of each diagnosis *d* given that result [24]. Applying Bayes’ theorem shows that *p*(*d*|*y*) depends on *p*(*y*|*d*)*p*(*d*), a factor that would need to be computed for separate diagnoses. Thus, for each diagnosis considered, a prior probability, or “pre-test suspicion”, [25] would be required [26], as well as the probability of the result given that diagnosis, estimable by measuring the frequency of that result among a sample of patients with the diagnosis.

Grouping all the diagnoses together, focusing only on “healthy” (*H*) versus “non-healthy” (*H*^*′*^), simplifies the matter. We need only compute

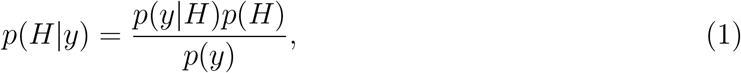

as illustrated in Fig. 1. (An intermediate simplification might be to group diagnoses into *D*, the set of diagnoses informed by the test, and *D*^*′*^, those that are not.) In Bayesian terms, we call *p*(*H*| *y*) the posterior, *p*(*y*|*H*) the likelihood, and *p*(*H*) the prior. Indeed, there is now only one prior to calculate: define criteria for who qualifies as healthy and ask what proportion of the population is healthy. Testing this healthy population will estimate *p*(*y*|*H*).

**Figure 1:**
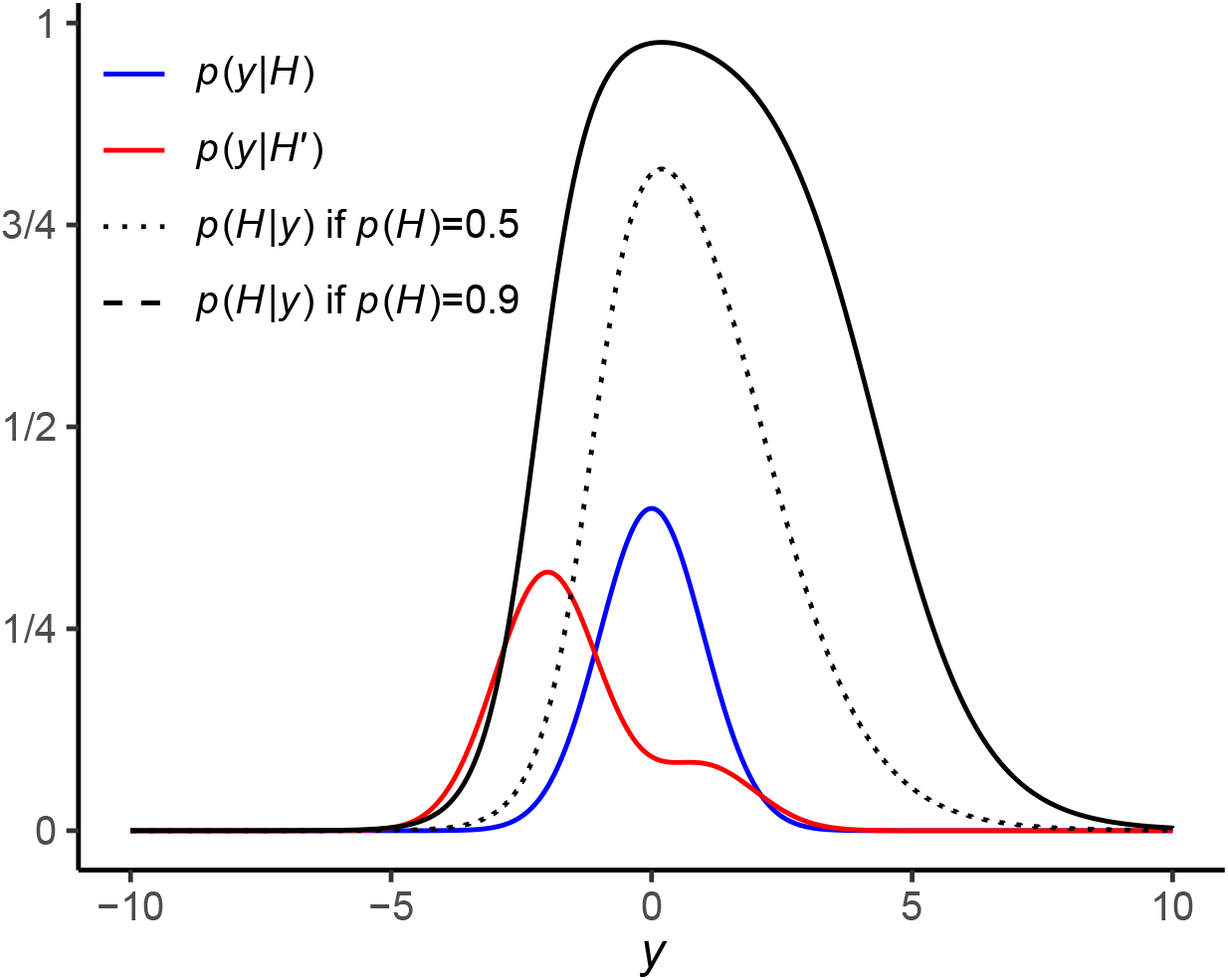
An example showing that the distribution of results among healthy and non-healthy people, combined with the prior probability of health, together influence the interpretation of a result with respect to health. Here, results among healthy people are normally distributed about zero, while results among non-healthy people are a mixture of two normal distributions on either side of the healthy mode. Due to the shape of the non-healthy distribution, results to the left of the healthy mode become concerning more quickly than to the right.

To sample

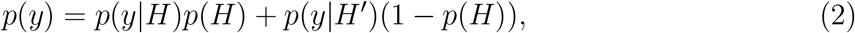

the overall distribution of results in the population, both healthy and non-healthy people should be tested. It is not only the distribution of results in the healthy population that is important, but also that of results among non-healthy people [2, 20, 21].

In practice, statistical interpretation of quantitative laboratory results is typically done with just *p*(*y*|*H*), what is called a (health-associated) reference distribution, representing results among apparently healthy patients [27]. Much has been written about how to empirically construct reference distributions. The standard procedure is that a sample of results from a healthy population is collected [3]. Then different methods, accounting for outliers [7, 28] and the shape of the resulting distribution, may be used to estimate quantiles of *p*(*y*|*H*) [9]. Parametric methods assume a form of the distribution while non-parametric methods are interested directly in the sample quantiles. To calculate the commonly used 95% interval, on the order of two hundred sample results are recommended [7, 8, 29].

Different studies could enforce different criteria for who is healthy, the definition of which can be controversial [30]. Some authors use a definition of health that seems aligned with computing *p*(*y*|*D*) [27, 31], Gräsbeck stating that “A person may be suitable as a healthy reference individual for one test … but unsuited for another,” while others use health criteria that are constant across tests [9], which aligns with computing *p*(*y*|*H*). We will use the *H* notation for simplicity, but in practice the particular criteria should be specified [3].

### Interpreting reference distributions for diagnosis requires assumptions about non-healthy results

The theoretical simplifications above result in an object, the reference distribution, that is relatively easy to estimate by sampling but does not give the full picture about the probabilities of different diagnoses given the patient’s test result [25]. An assumption that underpins current clinical use is that the extremeness of a result with respect to the reference distribution corresponds to how concerning it is. This is true, for example, if *p*(*y*|*H*^*′*^) is more diffuse than *p*(*y*|*H*) as in both limits of Fig. 1. But the distribution of results among non-healthy people can, in theory, flexibly change the implications of a patient’s result. If, to take an exceptional example, values of *y* among non-healthy people were for some reason concentrated at a high-probability interval of *y*| *H*, then extreme results would unintuitively be reassuring (Fig. S1A). And using Eq. 1 and Eq. 2, we can also see that extremes of an analyte unassociated with health (i.e. when *y*|*H* and *y*|*H*^*′*^ are identically distributed) would not be informative with respect to disease no matter how extreme *y* is with respect to the healthy population, although we should not expect this to ever be exactly the case especially since analytes are often chosen based on physiological connection with disease.

It is often implicitly assumed that extremes are only at the left and/or right limit of *y*. If true, the extremeness of the result can be quantified using the cumulative distribution function *F*(*y*|*H*), with high *F*(*y*|*H*), for example, corresponding to high extremes. Depending on the analyte, low extremes, high extremes, or both (as in Fig. 1) could be concerning. If both tails are concerning, one might quantify the extremeness of the result as, say, min (*F*(*y*| *H*), 1 − *F*(*y*| *H*)). Consider, however, a case in which there are two modes of *y* at which people are healthy, and non-healthy people also exhibit these two modes but more diffusely. (Perhaps they correspond to two modes of physical attributes, the sort of situation we analyze later in this paper.) Then concerning extremes are not exclusively at the left and right of the graph: they can also lie between the two modes (Fig. S1B). Directly applying a non-parametric method, using the rank order of reference results rather than their parametric form, to construct a reference interval would in this case obscure such nuances. It is sometimes held that non-parametric methods make no “assumptions as to the specific form of the underlying distribution of the data” [9] and that “they can be applied to any set of data, regardless of how the parent population of values is distributed” [31]. It is true that they can be applied as such, but as these cases show, there are assumptions in considering only the tails of the reference distribution as regions of concerning extremes.

Even if we know where the concerning extremes lie with respect to the reference distribution, there remains the question of how precisely to act on them. The risk of disease can vary continuously with the extremeness of the test result; there are often no clean boundaries on the result that dictate the occurrence of disease. But clinical decisions are often discrete: an action is taken or not. And so thresholds of extremeness, based on the available information, are set between courses of action. A 95% interval is typically used to define thresholds for concern [25, 32, 33], as it often is for other statistical analyses and illustrations (including in this paper). It is a convenient but ultimately arbitrary choice [25, 33–35] — why not use, say, 89% as a default for statistical analyses [36, ch. 4]? Ideally, rather, the probability of health combined with the costs and potential rewards of actions should dictate the optimal threshold [2, 21, 26, 29, 34, 37]. As Fig. 1 illustrates, the former depends on *p*(*y*|*H*^*′*^), as do the sensitivity and specificity of a threshold in detecting disease [21]. And indeed, a threshold on min (*F*(*y*|*H*), 1 − *F*(*y*|*H*)) may have different diagnostic ability according to the tail on which the extreme result lies.

### Jointly interpreting results depends on their relationship with each other (and with health)

Bayes’ theorem also allows us to interpret the levels of several analytes, say *y*_1_ and *y*_2_, together with respect to health:

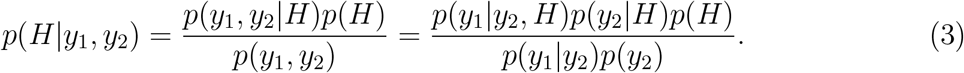

Conditioning on results of multiple tests would then seem to enable a more precise estimate of the probability of health, but when using solely reference distributions we again miss the non-healthy variation to exactly compute this probability. Thus it is not always advantageous to use the joint reference distribution of *y*_1_ and *y*_2_. We illustrate a variety of circumstances in Figs. 2 and S2.

**Figure 2:**
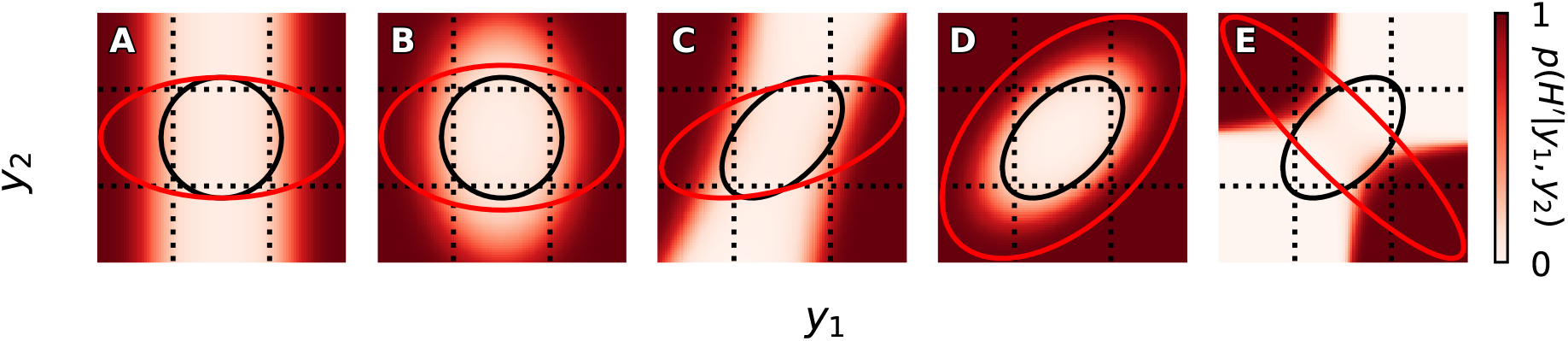
Examples of how the joint distributions of two test results *y*_1_ and *y*_2_ among healthy and non-healthy people are related to the posterior probability of health. Shown are 95% joint highest density regions of results among healthy (black ellipses) and non-healthy (red ellipses) people, as well as the 95% marginal highest density intervals among healthy people (dotted lines). In other words, the red ellipses contain 95% of the joint density of *y*_1_ and *y*_2_ in the non-healthy population, and the black regions represent joint reference regions as opposed to their univariate counterparts marked by dotted lines. The prior probability of being healthy is *p*(*H*) = 0.9 and *y*_1_ is associated with health, i.e. *y*_1_|*H* and *y*_1_|*H*^*′*^ are distributed differently, in every example. The analytes are independent in (A–B), and *y*_2_ is unassociated with health in (A) and (C).

The degrees to which *y*_1_ and *y*_2_ are each associated with health matter for whether to jointly consider them. For the sake of intuition, take the extreme case that *y*_2_ is distributed identically among healthy and non-healthy people, in other words that *y*_2_ is independent of health. This fact would be taken into account when computing the posterior, but not in the reference distribution *p*(*y*_1_, *y*_2_|*H*). In Figs. 2A and C, for example, extreme *y*_2_ is not in itself concerning, but the degree to which a particular *y*_1_ is extreme can be inappropriately altered by the inclusion of *y*_2_ [38]. It can thus be misleading to combine the reference distribution of *y*_1_ with *y*_2_, an effect alleviated as the association of *y*_2_ with health increases (Figs. 2B, D).

Whether it is advantageous to consider the joint distribution also depends on the relationship between *y*_1_ and *y*_2_. If *y*_1_ and *y*_2_ are independent among healthy people (Fig. 2A–B), the joint reference distribution *p*(*y*_1_, *y*_2_|*H*) simply separates into *p*(*y*_1_|*H*)*p*(*y*_2_|*H*), the product of the individual reference densities. The advantage of using joint reference distributions can be seen when *y*_1_ and *y*_2_ covary with a similar correlation among healthy and non-healthy people and are both associated with health; in Fig. 2D, the values of the posterior are diagonally symmetric and the reference distribution captures the shape. Even if *y*_1_ and *y*_2_ covary only among healthy people or non-healthy people, the posterior can follow a diagonal, sometimes behaving in ways unpredicted by the shape of the reference distribution (Fig. S2). The general advice that tests that are combined be “systematically” [39] or “physiologically” [40] related is sound: this implies that they covary. But these are not sufficient conditions because if they do not covary in a similar way between healthy and non-healthy populations, the reference distribution could be a poor proxy of the shape of the posterior distribution (Fig. 2E being an especially illustrative example).

The relationship between three blood analytes illustrates some of these concepts. The product of mean corpuscular hemoglobin (MCH) and red blood cell (RBC) count determines the hemoglobin concentration (HGB), a common measure of anemia as it quantifies how much oxygen a particular volume of blood can deliver. Based on this logic, MCH and RBC would better diagnose anemia together than apart, but is it better to use the reference distribution of HGB (a function of the MCH and RBC) or the joint distribution of MCH and RBC? The latter was observed in one study to be better diagnostic of mortality risk [15], suggesting it can be important to know whether MCH and RBC are jointly extreme even if HGB is not.

As others have noted, jointly considering analytes affects how one interprets the extremeness of a set of results. For example, a set of results that have no extremes apart may be extreme together [40] or vice-versa [41]. Proponents of joint reference regions have argued that the former effect makes the reference region more specific for interpretation [39]. Other proponents have argued that the latter effect controls for false positives due to multiple testing [41–43], while it has also been regretted that joint reference regions can bury the extremeness of a single test [40]. But as we have shown, the diagnostic impact of jointly considering analytes depends on their relationship with each other and with health. Joint reference distributions can be useful in some cases and misleading in others.

### Conditioning on features personalizes reference distributions

What if *y* depends on some random variable, a feature *x* unassociated with health, i.e. for which *x*|*H* and *x*|*H*^*′*^ are identically distributed? An implication of our analysis above is that jointly considering two covarying analytes where *y* is associated with health and *x* is not (as in Fig. 2C, taking *y* to be *y*_1_ and *x* to be *y*_2_), risks a misleading interpretation of the extremeness of *x* with respect to health. The joint reference distribution is *p*(*y, x*|*H*) = *p*(*y*|*x, H*)*p*(*x*|*H*).

But from Eq. 3 we see that *p*(*x*|*H*) = *p*(*x*|*H*^*′*^) implies

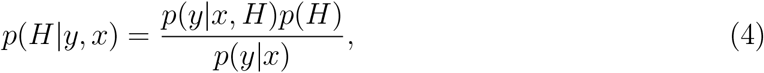

and so *p*(*x*|*H*) becomes irrelevant in the calculation of the posterior probability of health. The reference distribution that remains is *p*(*y*|*x, H*), which we can call the reference distribution of *y* specified to *x*.

This logic provides the theoretical justification for when the reference distribution of *x* can be disregarded. There may be other reasons to do so, including to avoid risks in interpreting joint distributions that we highlighted earlier, or simply to keep interpretation focused on *y*. It may be useful, for example, to measure the extremeness of a result adjusted to the patient’s age [44, 45] while not compounding this quantity with age itself as an indicator of concern; a 97-year-old’s result should not be flagged just because the patient is elderly.

However, *x* should not be disregarded with respect to the reference distribution of *y*. Suppose such a feature *x* exists: a feature of patients or their test conditions (e.g. the testing instrument or the time of day) not associated with health, or whose prevalence among healthy people one would like to disregard, but which is associated with the result *y*. Then while a random sample of the healthy population would exhibit the healthy variation in results in general [31], *p*(*y*|*H*), it might not estimate well the healthy variation *p*(*y*|*x, H*) one should expect given the patient’s feature [25]. Suppose the patient is an athlete and exercises far more than most people sampled. The greater the effect of exercise on healthy results, the more likely is a false positive for the patient when using a reference interval derived from the general healthy population. Creatine kinase, for example, is elevated under both dystrophy and after intense physical exercise [46].

Note that Eq. 4 shows that the interpretation of the reference distribution with respect to the probability of health may change depending on the features conditioned on. This would occur if, as features are conditioned on, the distribution of results among non-healthy people does not mirror changes in the reference distribution. Thus two results of the same analyte, under different feature values, which each fall outside their specified 95% reference interval may not necessitate the same level of concern [47]. (One consequence of this fact is that the argument that intra-person reference intervals, derived only from previous results from the patient, are preferable to inter-person ones [48] may not be so straightforward: how should thresholds be calibrated for each patient?)

With this caveat in mind, to adjust for a dichotomy in test results, perhaps if certain statistical criteria [10, 11, 22] indicate to do so, we might opt to consider only samples from people who share the differentiating feature, such as fellow athletes. This process is called subgrouping and it is the current practice for making reference intervals specific to certain features [3]. In reality, however, there are many features about the patient’s body and the conditions under which the test was performed [31, 49]. Progressively conditioning on each such *x* further limits the people who can be sampled and the test conditions under which they can be sampled.

It is also infeasible to collect a new reference sample specific to each patient. Instead, there might be data for a sample of patients, similar in some respects to the patient and dissimilar in others. Even if we were to subset the data to even two levels of specificity — say, to athletes over 25 years old — the number of samples with which to construct a more specific reference distribution would be substantially reduced. Framed in this way, it seems we must be satisfied with reference distributions in which we are confident (i.e. derived from a large reference sample) but which are perhaps not as specific as we might like.

### Reference distributions can be seen as posterior predictives of regression

We will see that regression provides a potential solution to this problem. Regression as a technique for constructing reference distributions is not new [50–52]. We will now motivate it and describe some of its principles from a theoretical perspective.

Let us begin by considering the problem of specifying reference distributions more formally. We have spent much of this paper discussing how to interpret reference distributions when they are exactly known. Let us adjust our notation to speak of inferring them from data. Let *X* be a matrix, the *i*th row of which comprises the *i*th reference individual’s values of the *k* features *x*_1_, …, *x*_*k*_, and let *y* be the vector of reference values, the *i*th element of which is the test result of the *i*th reference individual. We can call *X* and *y* together the reference data. Now let 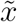 represent the features of the patient’s test (which was not part of the reference sample). A 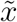 of the patient consisting of *k* = 3 features could be (1, 1, 0), possibly meaning: over 25 years old, athlete, not ambidextrous.

The reference distribution specified to the patient’s test is then the distribution of the result conditional on its features and the reference data: in probabilistic terms, 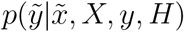. To infer a random variable’s relationship with features is to perform regression, and the distribution of new data based on observed data is called the posterior predictive. Thus, a reference distribution can be interpreted as the posterior predictive distribution of a regression.

To compute this distribution, it is necessary to specify a model that sets the form of the relationship between *X*_*i*_ and *y*_*i*_. A possible regression model is one that disregards the features and explains the result as simply a mean value *β*_0_ plus normal random error *ϵ*:

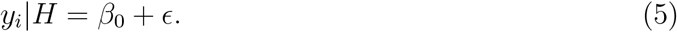

Reference distributions are usually constructed assuming this model, computing only 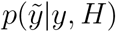. (Uncertainty in the parameters is ignored if 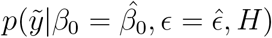 is computed, where 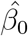 and 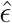 represent estimates of the parameters, e.g., when the reference distribution is assumed to be normal around the sample mean with variance equal to the sample variance [9].)

To calculate separate reference distributions for all combinations of *k* binary subgroups, it seems 2^*k*^ of these models must be created, *β*_0_ and *ϵ* inferred separately for each possible *X*_*i*_.

We again see our original problem; we would need to separately sample test results arising from 2^*k*^ combinations of features.

### With a good model, regression efficiently infers specific reference distributions

Note, however, that with this approach the effect of a feature is inferred anew for each combination of the remaining features; the effect of being an athlete would be inferred independently for younger and older patients. (We use “effects” to refer to regression coefficients; features need not be causal.) What if instead of modeling subgroups separately, we treated them as features which contribute, independently at least to some degree, to the result [22]? For example, in the form of a linear model in which

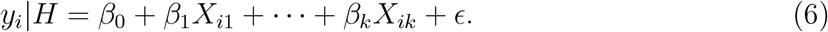

Returning to our example, this kind of regression infers the effect of being an athlete from both younger and older athletes, potentially extracting more value from the data [50].

This approach has risks. First, reference distributions can always be made more specific, increasing *k*, and at some level of specificity the data will be too limited to precisely infer feature effects. Uncertainty in the model parameters *θ* increases the variance of the posterior predictive distribution, which integrates over uncertainty in the parameters:

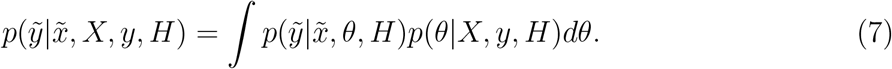

This equation reflects, for example, that if we are uncertain that the effect of a feature is as negative as it seems to be, then we should not be as surprised to see a large result than if we were certain in the effect. Well-considered, informative priors allow features to be incorporated into the model without unnecessarily increasing uncertainty in the reference distribution or overfitting the reference data, leading to more accurate reference intervals (Fig. 3). Setting priors such that in total they agree with the expected prior predictive variance [53, ch. 12] could also help; we should not expect variance in results to be arbitrary large or small.

**Figure 3:**
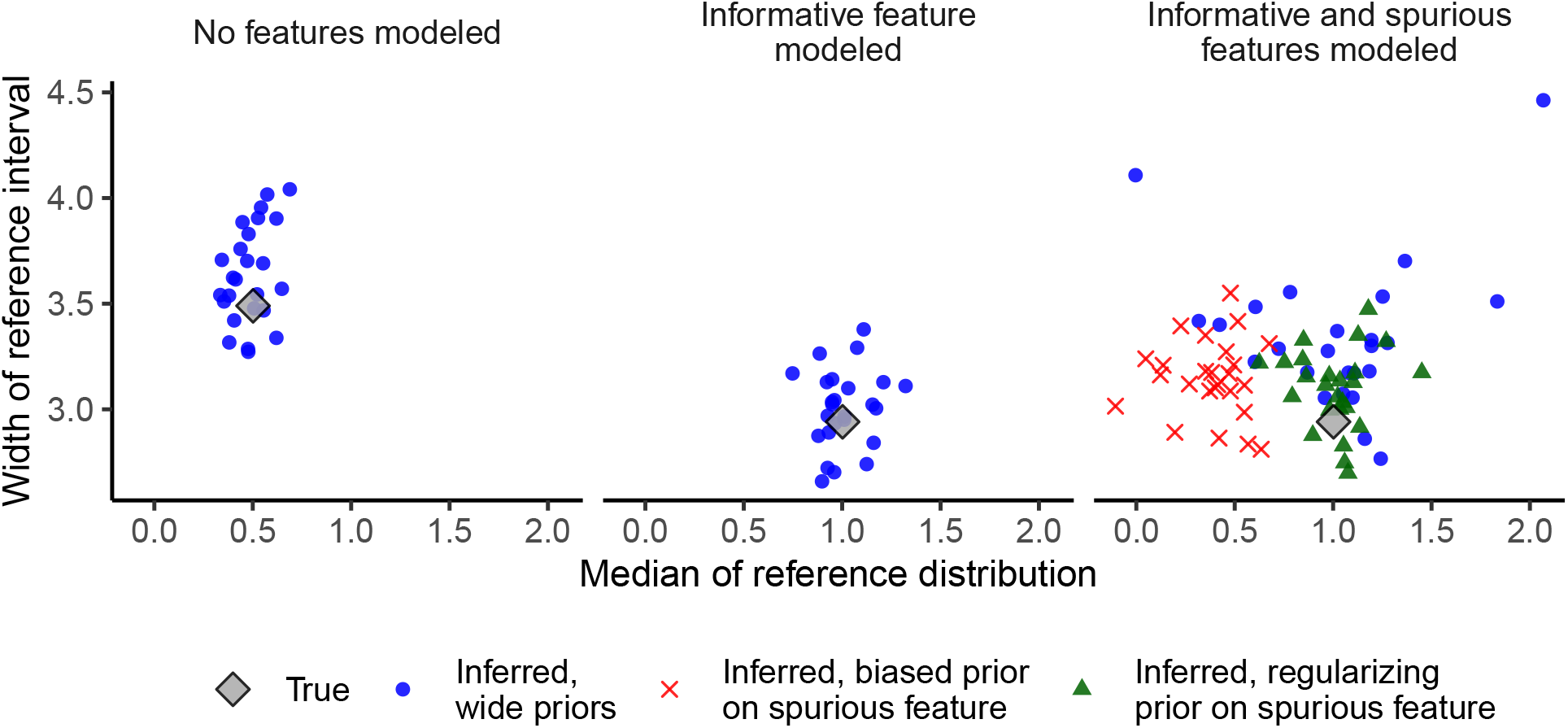
Inferring the 95% reference interval as two present binary features are progressively accounted for. Each point represents one reference interval, inferred from a sample of 100 simulated observations. One (informative) feature *x*_1_ occurs among 50% of people and its presence is associated with a unit increase in the result. The other (spurious) feature *x*_2_ occurs among 5% of people and has no association with the result. The model is *y* = *β*_0_+*β*_1_*x*_1_+*β*_2_*x*_2_+*ϵ* and the true coefficients are *β*_0_ = *β*_2_ = 0 and *β*_1_ = 1 with *ϵ* ∼ Normal(0, ¾). Different priors of the effect size *β*_2_ of the spurious feature are considered: Normal(0, 10) (wide prior), Normal(−1, ¼) (biased prior), and Normal(0, ¼) (regularizing prior). Including *x*_2_ should have no effect on the *x*_1_ = 1 posterior predictive since *β*_2_ = 0.

Second, models are always wrong to some degree, causing inaccuracies downstream in posterior predictive distributions. For example, we thus far assumed the variance of the error is constant, or homoscedastic. Suppose instead that error varies with some binary feature *x*_*j*_, lower when it is present and higher when absent. Fitting a homoscedastic model to this data, we may find that the error when *x*_*j*_ = 1 is overestimated and underestimated when *x*_*j*_ = 0, the posterior predictives being too wide and too narrow, respectively. The functional form of the feature effects may also be incorrect. This can occur, for example, when effects of some binary features *x*_*j*_ and *x*_*h*_ are modeled additively, but in fact their combined effect differs from their effects apart. This mis-specification skews coefficient estimates of the features that are accounted for (Fig. S3).

## Discussion

So how should we interpret a patient’s test result? Ideally, based on the results and prevalence of people with and without the diagnoses of interest, we could weigh the probability of the diagnoses. It is easier to consider just the likelihood that the result arose under healthy conditions, assuming the more extreme the result, the more concerning it is. A reference distribution, estimated from reference data of a sample of healthy people, provides these likelihoods. If other health-associated analytes covary with the one measured, it may, depending on the nature of the covariance, be useful to consider their joint reference distribution. Finally, it is possible that the effects of features recorded in the reference data could be inferred using a carefully specified regression model and be used to generate a posterior predictive distribution — a reference distribution that corresponds more specifically to the patient’s test.

In contrast to reference intervals, decision limits use clinical outcomes to define thresholds, thus taking healthy and non-healthy populations into account with respect to specific diagnoses. Decision limits are now used to interpret some tests [54, 55], but data to construct them is limited [29] and not many of them have been set [47]. It is generally agreed that they are preferable over their reference interval counterparts for interpreting tests [3, 29, 47, 56]. Our theoretical reasoning supports this conclusion; when the underlying healthy and non-healthy distributions and prior are exactly known, computation of the posterior probability of health (or a particular diagnosis) as in Eq. 1 is more precise and avoids the interpretive risks of relying solely on the reference distribution. (Likelihood ratios [57], e.g. *p*(*y*|*d*)*/p*(*y*|*d*^*′*^), capture one component of Eq. 1 but should be adjusted by the prior, e.g. *p*(*d*) [25].) Much of our reasoning about reference distributions would also apply to construction of decision limits.

We simulated univariate regression with respect to binary features to illustrate general considerations of using regression to specify reference distributions. With additional model assumptions, joint reference distributions could be derived by multivariate regression [58, ch. 16]. There are opportunities and risks in incorporating continuous features [59]. Assuming a functional form of the effect of a feature with respect to its value risks further model mis-specification. But if the function is correct, this approach has advantages over the traditional practice [45] of binning continuous features and sampling results independently within each bin. For one, intervals could be specified to the patient’s exact feature value, e.g. age rather than age bin. There can be additional complications in regression to the ones we showed, such as covariance between modeled features, which increases uncertainty about their effects [53, ch. 10] and thereby widens posterior predictives. We repeatedly assumed results are normally distributed for sake of demonstration, but this should not hastily be assumed in practice [60].

The Bayesian formulation of reference distributions could be applied to additional problems in constructing them. One such problem is reconciling differences between different laboratories [61, 62]. For example, multi-level modeling — which splits samples into groups, in each of which parameter estimates are partially pooled with those of other groups [63] — could be used to capture the dependence of effects on the laboratory. Current practice in transference of reference intervals between laboratories, important for reducing costs, is through validation: using a smaller number of samples than recommended for constructing an interval from scratch, validate that an existing interval applies to the new laboratory, and use it if it does [3]. Although intervals can align well with one another [12], there is a dead end in the procedure if they do not. Multi-level modeling would allow data from other laboratories to be used to reduce the sample burden even if intervals are not strictly consistent.

Overall, a theme of our work has been the importance of the marginal likelihood, *p*(*y*), or in the case of regression, *p*(*y*|*x*), in interpreting reference distributions. Bayes’ theorem uses it directly. If it is not available, however, reference distributions can be used, provided the assumptions on the missing factors are acknowledged. Regression can be applied to take more and more features into account, gradually personalizing interpretation of results from limited data. Fortunately, there is much wisdom about Bayesian modeling and regression [36,53,58,63]. Perhaps by further tracking technical and physiological details of tests [26], sharing our ever-increasing data [64], and iterating through careful modeling and criticism [65], we will be able to realize clinical laboratory diagnostics with growing precision.

## Methods

Linear regression was performed using rstanarm [66], built on the Stan programming language [67]. We set a Normal(0, 10) prior over coefficients unless otherwise stated, and a Exponential(1) prior over the standard deviation of the error *ϵ*. Code can be accessed at https://github.com/berkalpay/reference-distributions.

## Data Availability

Code can be accessed at https://github.com/berkalpay/reference-distributions.

## Acknowledgements

We thank Rachel Petherbridge, Thomas Dupic, and Derek Aguiar for useful discussions and comments. B.A.A. was supported by an NSF Graduate Research Fellowship under Grant No. DGE-2140743.

