## Supplementary material for "Some statistical theory for interpreting reference distributions"

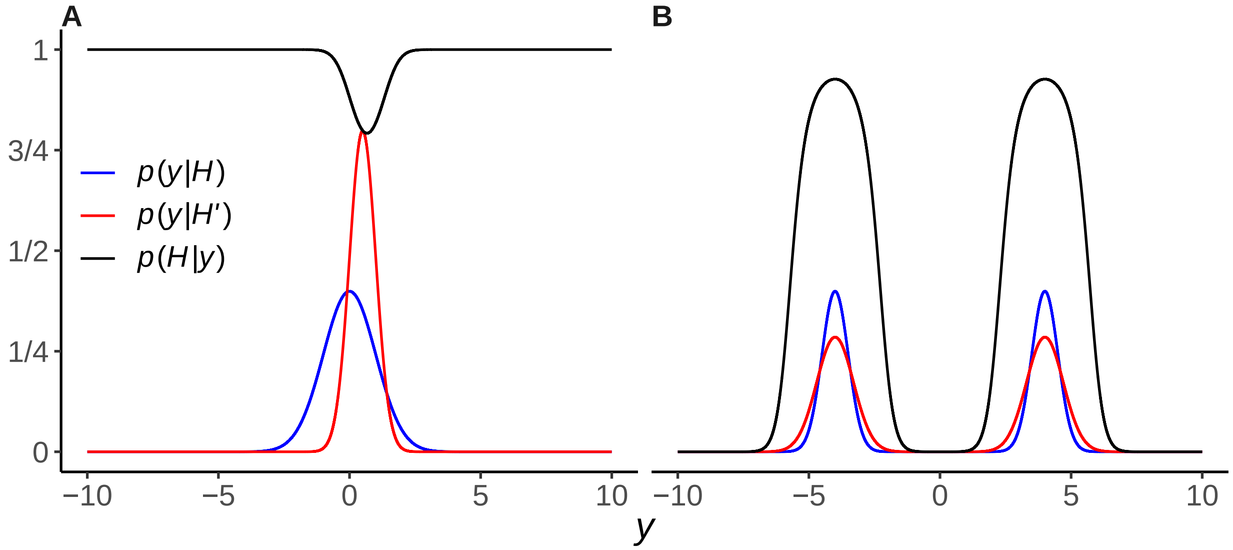


**Figure S1:** More examples as in Fig. 1.

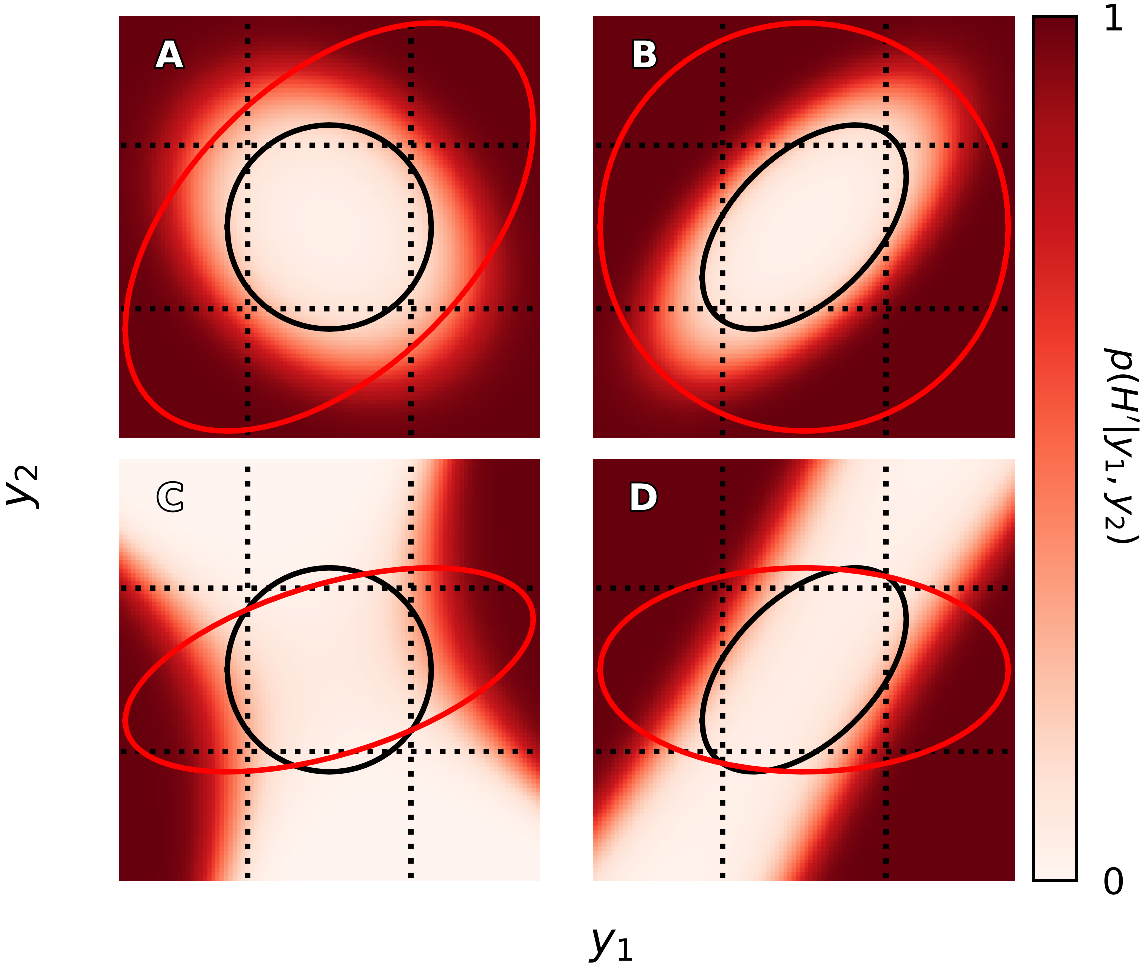


**Figure S2:** More examples as in Fig. 2, focusing on when $y_1$ and $y_2$ covary among healthy people or non-healthy people but not both.

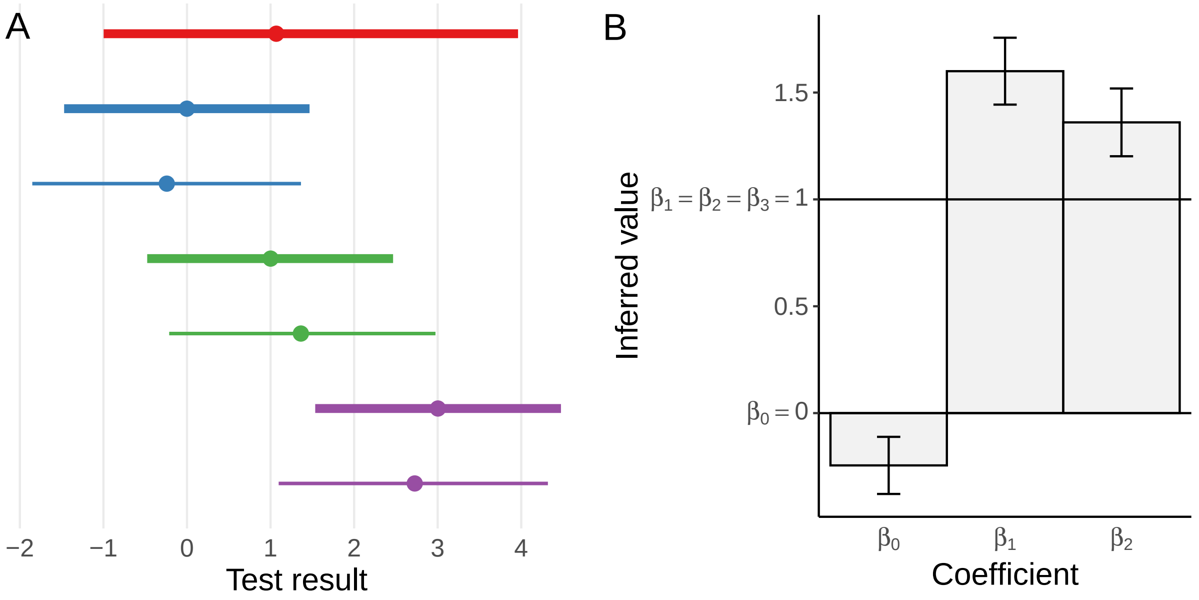


**Figure S3:** Inferences of a linear model of test results from 100 simulated observations when an interaction term is unaccounted for. There are two binary features, $x_1$ and $x_2$, both with 50% prevalence. The model is $y = \beta_0 + \beta_1 x_1 + \beta_2 x_2 + \epsilon$ but there is in truth an additional interaction term $\beta_3 x_1 x_2$. The true coefficients are $\beta_0 = 0$ and $\beta_1 = \beta_2 = \beta_3 = 1$, and the error is $\epsilon \sim \text{Normal}(0, {}^3\!/\!_4)$. (A) 95% central intervals of true (bold lines) and posterior predictive distributions (thin lines) of results in the population (red) as features are conditioned on: $x_1 = x_2 = 0$ (blue), $x_1 = 1$ and $x_2 = 0$ (green), and $x_1 = x_2 = 1$ (purple). Points represent medians. (B) Mean posterior coefficient estimates with standard errors.
